# Resident Physician Selection Practices and Professionalism-Related Difficulties in Japan: A Nationwide Cross-sectional Survey

**DOI:** 10.64898/2026.08.05.26359757

**Authors:** Miwa Sekine, Yuji Nishizaki, Takashi Watari, Kiyoshi Shikino, Sho Fukui, Kazuya Nagasaki, Masanori Nojima, Taro Shimizu, Yu Yamamoto, Hiroyuki Kobayashi, Yasuharu Tokuda

**Affiliations:** Division of Medical Education, Juntendo University Faculty of Medicine, Tokyo, Japan; Integrated Clinical Education Center, Kyoto University Hospital, Kyoto, Japan; Department of Community-Oriented Medical Education, Chiba University Graduate School of Medicine, Chiba, Japan; Department of Emergency and General Medicine, Kyorin University, Tokyo, Japan; Department of General Medicine, Fujita Health University, Aichi, Japan; Center for Translational Research, The Institute of Medical Science, The University of Tokyo, Tokyo, Japan; Department of Diagnostic and Generalist Medicine, Dokkyo Medical University Hospital, Tochigi, Japan; Division of General Medicine, Center for Community Medicine, Jichi Medical University, Tochigi, Japan; Department of Internal Medicine, Mito Kyodo General Hospital, University of Tsukuba, Ibaraki, Japan; Muribushi Okinawa Center for Teaching Hospitals, Okinawa, Japan; Tokyo Foundation for Policy Research, Tokyo, Japan; Clinical Translational Science, Juntendo University Graduate School of Medicine, Tokyo, Japan

**Keywords:** resident selection, graduate medical education, professionalism, structured interviews, Japan

## Abstract

**Introduction:** Postgraduate clinical training is crucial for developing professional competence, communication skills, and effective teamwork. Although resident physician selection is crucial, little is known about how Japanese residency programs select residents and whether selection practices are associated with difficulties during training.

**Methods:** We conducted a nationwide cross-sectional survey of residency programs participating in Japan’s 2023 General Medicine In-Training Examination (GM-ITE). Program directors completed a questionnaire assessing selection methods, interview content, quality-assurance measures, and resident difficulties, defined as at least one postgraduate year 1 or 2 resident physician receiving disciplinary action or a severe warning. Free-text responses were coded using the Situation, Task, Action, and Result framework. Associations between selection methods and resident difficulties were examined using adjusted logistic regression models controlling for hospital type and number of GM-ITE examinees.

**Results:** Of 151 participating physician-selection programs, 150 provided valid responses. Interviews were used by 90.1% of programs and were identified as the most important selection component by 87.3%. Thirty-five programs (23.3%) reported difficulties with resident physicians, involving professionalism and workplace conduct including rule, ethics, or boundary violations, work avoidance or unavailability, and inappropriate communication. Use of applicants’ pre-clinical-clerkship computer-based test scores as a selection criterion was associated with resident difficulties (adjusted OR, 4.60; 95% CI, 1.50–14.11; P = 0.008; FDR-adjusted P = 0.048). No significant associations were observed for essays, academic tests, medical school grades, or personality assessment. Program-level GM-ITE total and domain scores did not differ significantly between programs with and without reported resident physician difficulties.

**Discussion:** Resident physician selection in Japan is highly interview-centered; reported difficulties were more often related to professionalism and workplace conduct than to knowledge deficits. Although these exploratory findings are program-level, they highlight the importance of strengthening the quality assurance processes in resident selection systems, particularly for assessing professionalism-related attributes in applicants.

**Practice Points:**

- Resident selection in Japan is strongly interview-centered; however, explicitly structured interviews, scoring systems, and rater calibration remain uncommon.
- Program-reported resident difficulties primarily involved professionalism, communication, reliability, and workplace conduct rather than knowledge deficits.
- Academic selection indicators alone may be insufficient to identify applicants who later require professionalism-related support.
- Residency programs should strengthen interview quality assurance through clearer structure, explicit evaluation criteria, and interviewer calibration.
- Selection should be complemented by early identification, feedback, and longitudinal support during residency.

## Introduction

Postgraduate clinical training is a formative period during which newly graduated physicians develop the foundation of clinical knowledge, professional attitude, communication skills, and readiness to work within clinical teams. Given that this period shapes trainee-physicians into practicing professionals, the process through which trainees enter postgraduate training requires careful attention. Therefore, residency selection is not simply an academic ranking process, but a multidimensional assessment process that may include academic metrics, interviews, motivation, professionalism, communication, and fit with the training program [1–3].

In Japan, newly graduated physicians enter a mandatory two-year basic postgraduate clinical training program after medical licensure [4]. In this study, “resident physician” refers to physicians in this two-year basic postgraduate training program, rather than specialty trainees in later-stage residency programs. This program is designed as a broad-rotation training system in which resident physicians rotate across multiple departments over a two-year period. This period is critical for developing the foundation for professional clinical practice. Residency training develops not only resident physicians’ clinical knowledge and practical skills but also their professional attitudes and behaviors. Therefore, the process of selecting incoming physicians by residency programs deserves careful attention for comprehensive physician development. Globally, residency selection is not simply an academic ranking process but a multidimensional assessment process that combines academic and non-academic information. For example, studies from the United States, Canada, and Jordan describe the use of combinations of academic screening, standardized examinations, interviews, letters of recommendation, and ranking procedures; however, the specific components and their relative weights vary across settings and specialties [5–7]. Moreover, many selection processes attempt to assess attributes that are difficult to capture through knowledge tests alone, including motivation, communication, professionalism, program fit, and readiness to work within clinical teams. However, the extent to which these approaches are used, structured, or quality assured may differ substantially across training systems.

Resident selection in Japan also involves multiple components, including interviews, written examinations, medical school grades, and computer-based testing scores. The selection processes are designed to identify applicants who align with each institution’s educational goals, values, and desired resident characteristics and thereby also reflect the distinctive features of the training institutions. Previous studies in Japan have examined specialty preferences, residency-matching systems, applicants’ hospital preferences, the geographic maldistribution of residents, and the effects of postgraduate rotation programs on clinical knowledge [8–13]. However, these studies have focused mainly on resident choice, matching outcomes, geographic distribution, or training outcomes, rather than on how residency programs select incoming residents. The selection practices used in Japanese residency programs, including interview content and approaches to quality assurance, remain underexplored. Furthermore, it remains unclear whether specific selection practices are associated with program-reported resident physician difficulties or program-level clinical knowledge during residency.

To explore resident selection practices in Japan, this study aimed to (1) describe how Japanese residency programs select incoming residents and (2) examine whether these practices were associated with program-reported resident physician difficulties and program-level GM-ITE performance. We conducted a nationwide cross-sectional survey and analyzed the data for associations using adjusted regression models accounting for hospital type and the number of GM-ITE examinees.

## Methods

### Participants

We conducted a cross-sectional study of Japanese postgraduate residency training programs in hospitals administering the 2023 General Medicine In-Training Examination (GM-ITE). The GM-ITE is a voluntary annual examination for resident physicians in Japan. The target respondents were program directors or supervising physicians responsible for postgraduate years 1 and 2 (PGY1 and PGY2, respectively) residency training at the participating institutions. Because this study focused on program-level selection practices and program-level outcomes, the unit of analysis was the training program or the hospital. A total of 151 programs were included in this study.

### Recruitment and ethical considerations

Recruitment was integrated into the GM-ITE process and facilitated by the Specified Nonprofit Corporation, Japan Institute for Advancement of Medical Education Program (JAMEP). This study was approved by the ethical review board of JAMEP (No 23-28). The study was reported according to the Strengthening the Reporting of Observational Studies in Epidemiology guidelines. Informed consent for the use of the questionnaire responses was obtained from the respondents through an online checkbox before questionnaire completion. For the use of linked program-level GM-ITE and training environment survey data, study information was disclosed on the JAMEP website and handled through an opt-out process. The participants were informed that the study involved no medical intervention and that no disadvantage would result from non-participation. Data confidentiality was ensured through anonymization and secure data management procedures. This study was conducted in accordance with the principles of the Declaration of Helsinki.

### Questionnaire and study variables

Questionnaire data were collected from eligible respondents at institutions participating in the GM-ITE in the fiscal year 2023. The GM-ITE is a computer-based formative assessment administered to Japanese resident physicians to evaluate achievement during postgraduate clinical training and to support training guidance and program improvement. In this study, the program-level average GM-ITE total and domain scores were used as secondary program-level outcomes. Domain scores included general principles (medical interview/professionalism), symptomatology/clinical reasoning, physical examination/clinical skills, and disease-specific theories. The questionnaire assessed resident selection practices, interview content, interview quality assurance measures, and program-reported difficulties faced by the resident physicians. Because the questionnaire asked whether such cases had ever occurred, this outcome was interpreted as a program-level report of prior experience rather than an incident outcome temporally linked to the 2023 selection process. The English translation of the questionnaire is provided in Supplementary Appendix 1. For selection practices, respondents were asked what methods they employed in their hospital’s resident selection examination, including interviews, essays, written academic tests, medical school grades, computer-based test (CBT) scores, and aptitude tests.

In Japan, CBT is part of the Common Achievement Tests administered by the Common Achievement Tests Organization (CATO), which has been established as a nationwide organization of medical and dental schools. The CBT is a computer-based standardized knowledge assessment conducted before clinical clerkship [4]. Medical students are required to pass this examination before entering a clinical clerkship, as it ensures that they are qualified to participate in patient care under supervision. CBT questions were drawn from a large item pool developed and reviewed by CATO, and scoring was based on Item Response Theory. In this study, the CBT score was treated as a standardized knowledge-based selection indicator.

For assessing resident physicians’ difficulties reported during the training program, respondents were asked whether any PGY1 or PGY2 resident at their institution had ever received a disciplinary action or a severe warning. Respondents who reported difficulty with resident physicians were asked to describe the most concerning reported case using a structured free-text format based on the Situation, Task, Action, and Result (STAR) framework. This framework is a practical format commonly used in behavior-based interviews to elicit specific descriptions of past behavior in context, including the situation or task, action taken, and result [14]. This approach is consistent with structured behavioral interviewing, which emphasizes specific descriptions of past behavior rather than global impressions or unstructured judgments [15]. Facility-level background variables were compiled from hospital websites and external sources rather than from the questionnaire itself. Questionnaire responses were linked to the facility-level GM-ITE data and training environment survey data using anonymized institutional identifiers. Information on reported resident physician difficulties when described in the free-text responses was additionally coded.

### Coding of open-ended responses

Open-ended responses to important interview questions, interview quality assurance measures, and program-reported difficulties of resident physicians were coded using an inductive content coding approach. For the interview-related questions, the responses were reviewed and grouped into thematic domains based on their substantive meanings. Multiple themes were assigned to a single response when multiple concepts were presented. For analyzing resident physician difficulties, free-text descriptions based on the STAR framework were coded separately for the situation, task, action, and result for each past behavior.

ChatGPT (primarily GPT-5.4 Thinking; OpenAI) was used to support the preliminary identification of candidate concepts from free-text responses. It was also used to assist with English-language refinement of the manuscript. AI-generated suggestions were reviewed and revised by the authors, and final coding decisions and interpretations were made exclusively by the investigators. One investigator reviewed the responses, developed the initial coding scheme, and assigned codes. The coding scheme and assigned codes were then reviewed by multiple investigators, and ambiguous cases and coding discrepancies were discussed until a consensus was reached. Codes were assigned at the program level, with one reported case representing each program. When responses contained multiple concepts, coding was based on the explicit content of the response, without inferring information that was not stated.

### Statistical analysis

All analyses were conducted at the program level. Categorical variables are summarized as counts and percentages, and continuous variables are summarized as means with standard deviations, medians with interquartile ranges, and ranges, as appropriate. For comparisons between programs with and without resident physician difficulties, categorical variables were compared using the chi-square test, Fisher’s exact test, or chi-square test with simulated p-values when appropriate for sparse tables. Continuous variables were compared using the Wilcoxon rank-sum test, except for program-level GM-ITE total and domain scores, which were compared using Student’s t-test.

The primary dependent variable was the presence of program-reported resident physician difficulty. Because the questionnaire asked whether such cases had ever occurred, this outcome was interpreted as a program-level report of prior experience rather than an incident outcome temporally linked to the 2023 selection process. In the analytic coding, programs were classified as having resident physician difficulty if either disciplinary action or severe warning was reported, and as having no resident physician difficulties if no applicable resident was reported. Programs with missing responses to this item were treated as missing and excluded from analyses involving the resident physicians’ difficulty outcome.

To examine the associations between selection practices and the presence of program-reported resident physician difficulty, adjusted logistic regression models were fitted, with one selection practice variable entered at a time. Common adjustment variables were hospital type and the number of GM-ITE examinees; the number of GM-ITE examinees was used as a proxy for resident physician cohort size, which may be related to the probability of observing at least one program-reported resident physician difficulty. The selected non-interview selection methods examined in the main analyses included essays, written academic tests, medical school grades, CBT scores, and aptitude tests (personality and ability assessments). As a supplementary analysis, a grouped knowledge-based selection indicator was created, defined as the use of a written academic test conducted by the program and/or applicants’ pre-clinical clerkship CBT scores as selection criteria. This grouped indicator was examined in relation to the presence of program-reported difficulties for resident physicians. For secondary program-level outcomes, descriptive comparisons of GM-ITE total and domain scores were conducted according to the presence of program-reported resident physician difficulty. In the supplementary analyses, associations between selection practices and program-level GM-ITE total scores were examined using separate linear regression models adjusted for hospital type and number of GM-ITE examinees.

For the descriptive analysis of free-text resident physician difficulty descriptions, STAR-coded features were summarized among programs reporting a resident physician’s difficulty; because multiple codes could be assigned to a single case, percentages were not mutually exclusive. For the open-ended responses to important interview questions and interview quality assurance measures, the response contents were grouped into thematic domains and themes for descriptive analysis. Multiple themes were assigned to a single response; therefore, the percentages were not mutually exclusive. False discovery rate-adjusted p-values were calculated using the Benjamini– Hochberg method for regression analyses to examine the associations between the selection methods and study outcomes. The P-values in the descriptive comparison tables were not adjusted and were interpreted as exploratory. Missing data were handled using complete-case analysis for each model. The data were analyzed using Microsoft Excel, and statistical analyses were performed using R version 4.5.1.

## Results

### Program characteristics and selection practices

A total of 151 residency programs were included in the analysis. Most of the respondents were program directors (84.1%). Most programs were based in community hospitals (90.7%), and 64.9% were in rural areas. The median number of GM-ITE examinees per program was 13.0 (IQR, 7.5–22.0).

Interview-based selection predominated: 90.1% of the programs used individual in-person interviews, and 87.3% identified the interview as the most important selection component. Among non-interview methods, essays (50.3%) and medical school grades (37.7%) were relatively common, whereas written academic tests (16.6%), CBT scores (11.3%), and aptitude tests were less frequently used. Detailed program characteristics and selection practices are provided in Table 1.

**Table 1.**
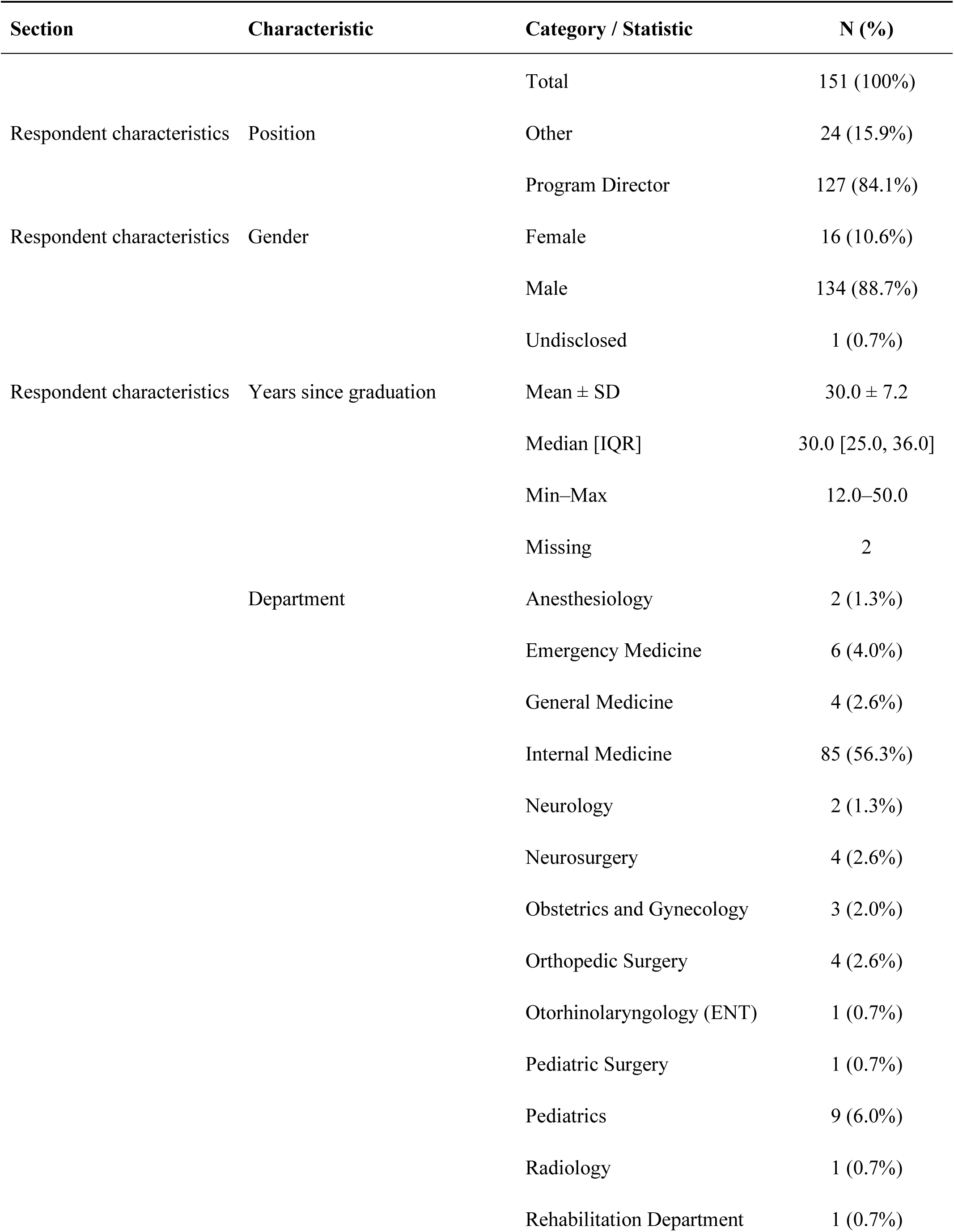
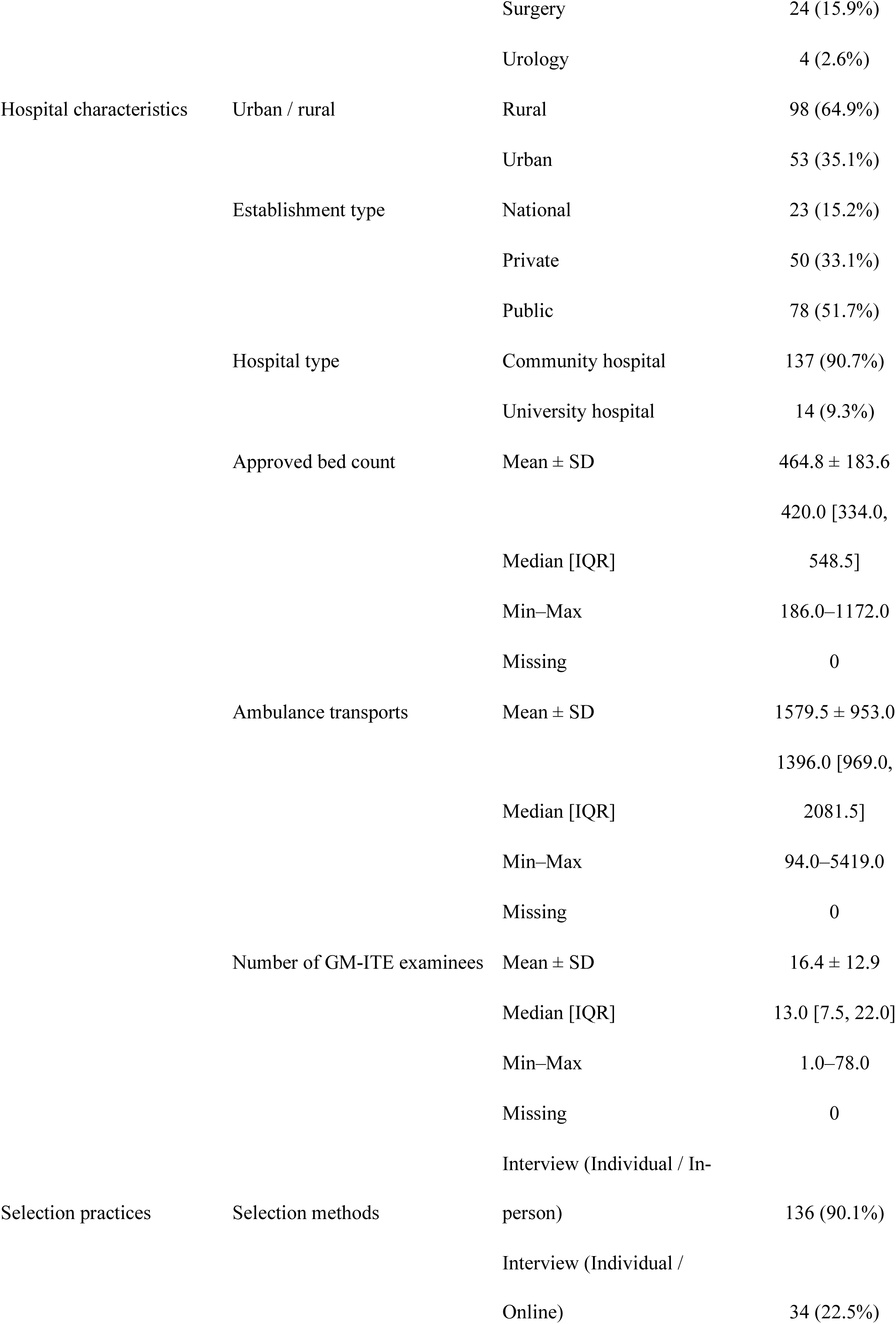
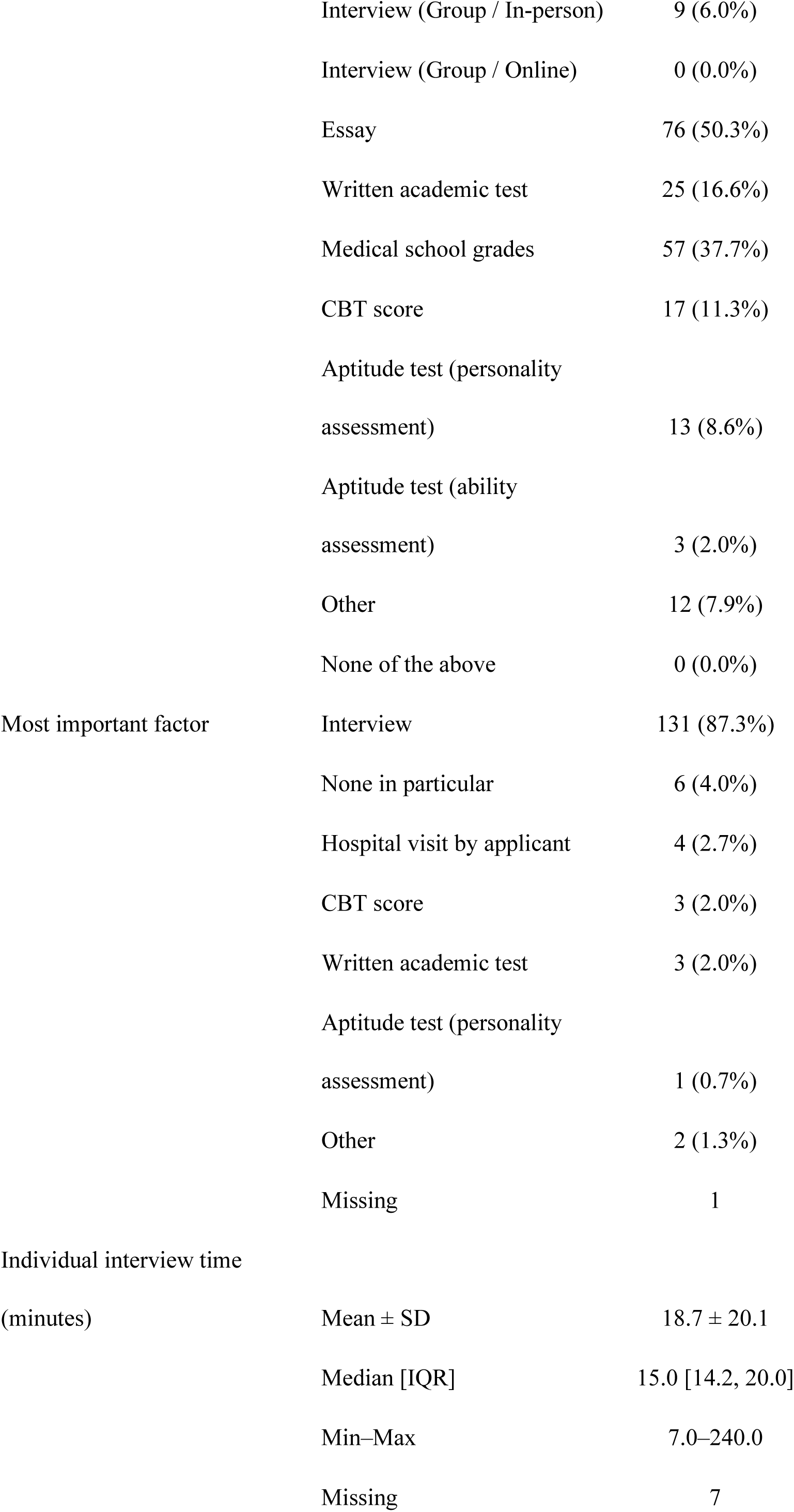
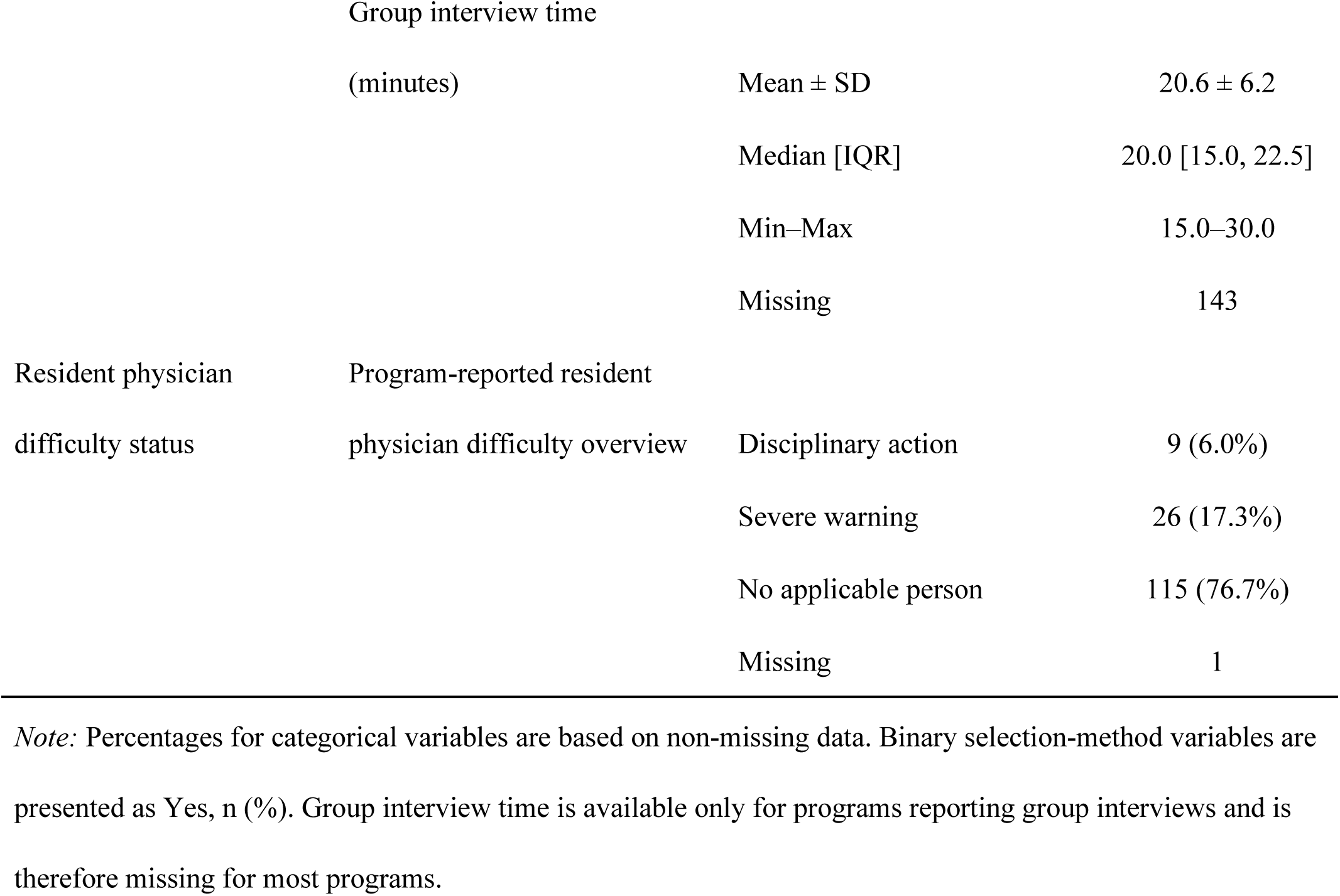
Respondent characteristics, program characteristics, resident physician selection practices, and resident physician difficulty status.

Among the 151 programs, 150 provided valid responses regarding the resident physicians’ difficulty statuses. Of these, 9 (6.0%) reported having at least one resident who received disciplinary action, and 26 (17.3%) reported having at least one resident who received a severe warning. Programs were classified as having program-reported resident physician difficulty if either a disciplinary action or a severe warning was reported, yielding 35 out of 150 programs (23.3%).

Program characteristics were broadly similar between programs with and without reported difficulty for resident physicians, although programs with reported difficulty tended to have a larger cohort of resident physicians. The use of CBT scores was more common among programs with resident physician difficulty than among those without such difficulties (28.6% vs. 6.1%, P < 0.001). No clear differences were observed for the other non-interview selection methods or individual interview times. The program-level GM-ITE total and domain scores did not differ significantly between the two groups. Detailed comparisons by the resident physicians’ difficulty status are presented in Table 2.

**Table 2.**
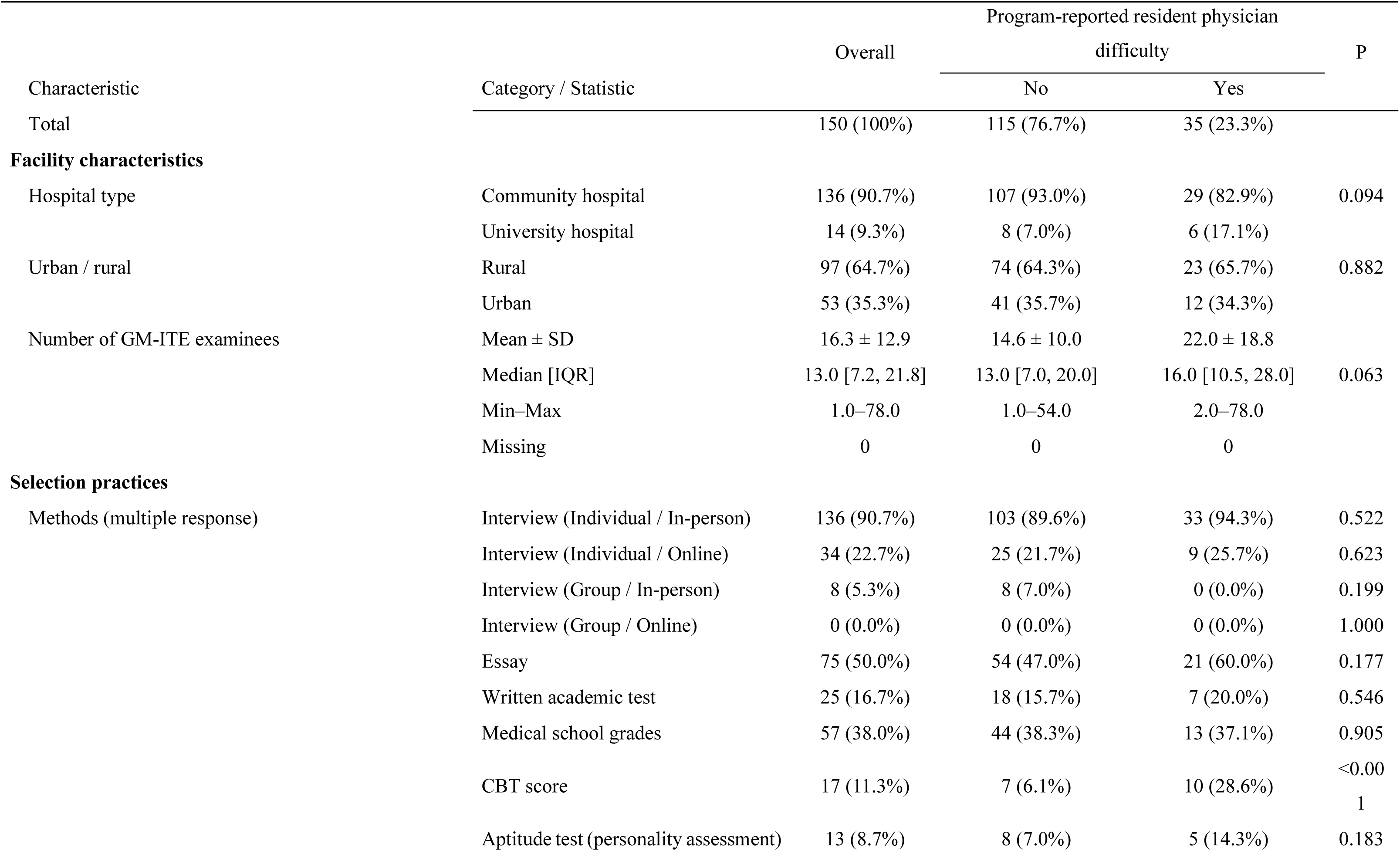
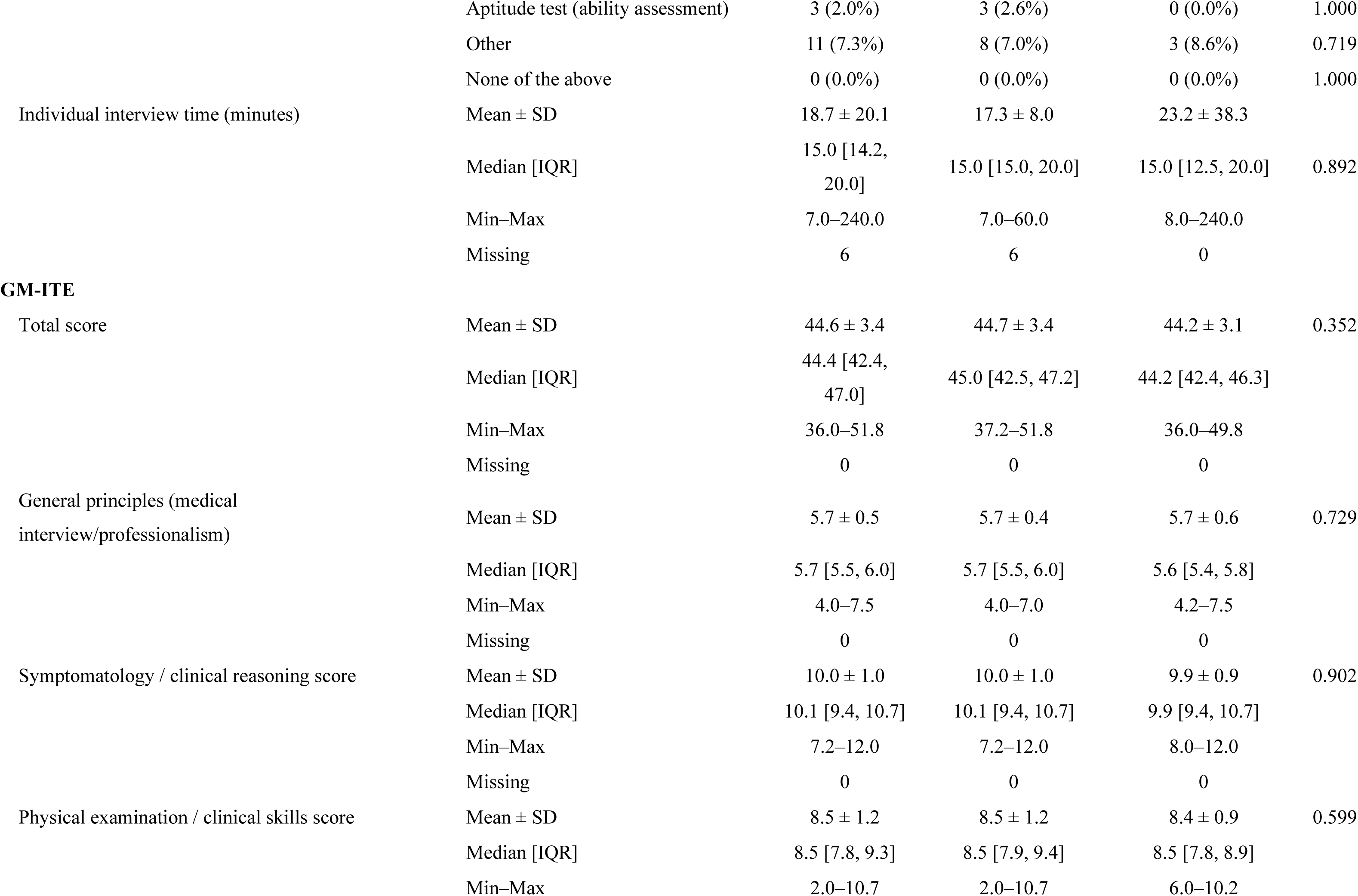
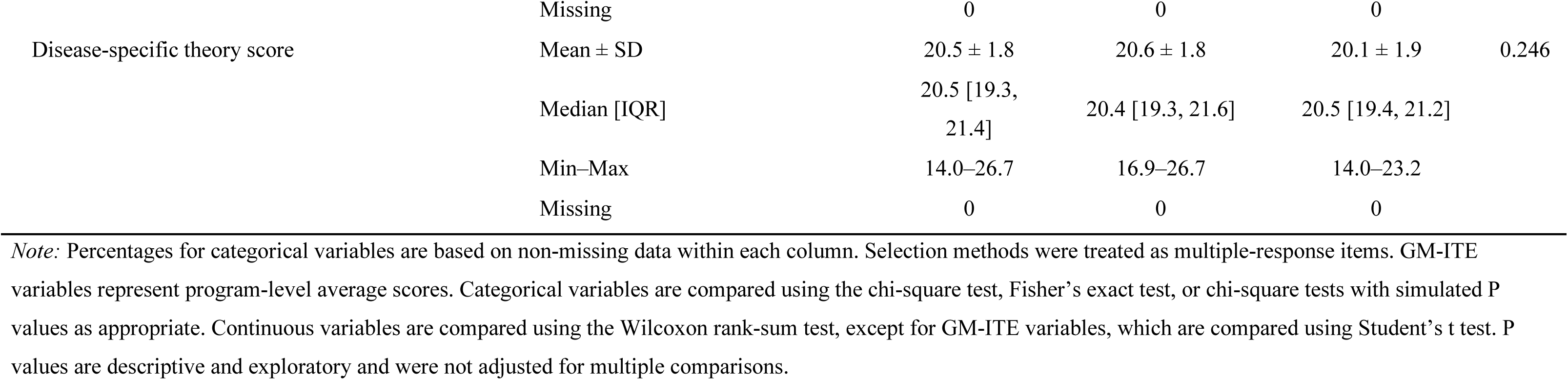
Program characteristics, resident physician selection practices, and resident outcomes based on the presence of program-reported resident physician difficulty.

### Interview question domains and interview quality-assurance practices

Because interviews were the predominant selection method, we further summarized the content of the most important interview questions and the measures used to ensure interview quality. Among the open-ended responses to the most important interview questions, the most frequently identified themes were: “why this hospital/program” (63 programs, 41.7%) and “career plans” (53 programs, 35.1%). Other commonly reported themes included “student activities/life history” (29 programs, 19.2%), “program fit or what the applicant was seeking” (29 programs, 19.2%), and “why medicine” (23 programs, 15.2%). Themes related to “teamwork and inter-professional collaboration” (14 programs, 9.3%), “attitude/professionalism” (13 programs, 8.6%), and “motivation/enthusiasm” (12 programs, 7.9%) were also reported. Overall, interview questions tended to focus more on applicants’ motivation, future orientation, and fit with the program than academic knowledge alone.

The most frequently reported measure used to ensure interview quality was a multiple-interviewer system (127 programs, 84.1%), followed by the inclusion of non-physician interviewers (50 programs, 33.1%). Less frequently reported measures included scoring/quantification (12 programs, 7.9%), bias control/adjustment (10 programs, 6.6%), structured interviews (9 programs, 6.0%), and multistage or additional selection methods (7 programs, 4.6%). Eighteen programs (11.9%) did not describe specific quality-assurance measures. Detailed themes are listed in Table 3.

**Table 3.** Themes identified from the open-ended responses to important interview questions and measures to ensure interview quality.

**Panel A. Important interview questions**
| Domain | Theme | Mentioned, n (%) |
| --- | --- | --- |
| Academic/clinical orientation | Medical interest / curiosity | 2 (1.3%) |
| Background and experiences | Student activities / life history | 29 (19.2%) |
| Career vision | Career plan | 53 (35.1%) |
|  | Ideal doctor / professional identity | 10 (6.6%) |
|  | Specialty preference | 10 (6.6%) |
|  | Geographic plan | 7 (4.6%) |
|  | Research career | 1 (0.7%) |
| Communication style | Self-presentation | 2 (1.3%) |
| Interpersonal competencies | Teamwork / interprofessional | 14 (9.3%) |
|  | Communication / conflict | 11 (7.3%) |
| Leadership and teamwork | Leadership / coordination | 3 (2.0%) |
| Learning orientation | Reflection / learning from experience | 4 (2.6%) |
| Motivation and purpose | Why medicine | 23 (15.2%) |
|  | Motivation / enthusiasm | 12 (7.9%) |
| Personal attributes | Resilience / stress tolerance | 7 (4.6%) |
|  | Adaptability / flexibility | 2 (1.3%) |
|  | Independence / autonomy | 1 (0.7%) |
| Policy/other | Others | 13 (8.6%) |
|  | Holistic evaluation policy | 1 (0.7%) |
| Professionalism | Attitude / professionalism | 13 (8.6%) |
| Program fit | Program fit / what they want | 29 (19.2%) |
|  | Training priorities | 11 (7.3%) |
| Program selection | Why this hospital / program | 63 (41.7%) |
|  | Intention to stay at this hospital | 3 (2.0%) |
|  | Comparison with other hospitals | 2 (1.3%) |
|  | Identify weaknesses / critical thinking | 2 (1.3%) |
| Thinking and execution | Problem solving / PDCA | 3 (2.0%) |

**Panel B. Measures to ensure interview quality**
| Domain | Theme | Mentioned, n (%) |
| --- | --- | --- |
| Evaluation process | Scoring / quantification | 12 (7.9%) |
|  | Use of non-interview information | 5 (3.3%) |
| Interview environment | Creating a relaxed environment | 4 (2.6%) |
| Interview structure | Multi-interviewer system | 127 (84.1%) |
|  | Inclusion of non-physicians | 50 (33.1%) |
|  | Structured interview | 9 (6.0%) |
| Non-response / no specific measure | No specific measure described | 18 (11.9%) |
| Quality assurance | Bias control / adjustment | 10 (6.6%) |
|  | Rater calibration | 4 (2.6%) |
| Quality improvement | Feedback and reflection | 2 (1.3%) |
| Selection design | Multi-stage or additional selection methods | 7 (4.6%) |
| Other | Others | 4 (2.6%) |

*Note:* Themes were derived from open-ended responses regarding the most important interview questions and measures used to ensure interview quality. Multiple themes could be assigned to a single response; therefore, percentages are not mutually exclusive.

### Content of program-reported resident physician difficulties based on the STAR framework

In the Situation domain, resident physician difficulties were reported slightly more often in on-duty clinical or training settings than in off-duty or private settings (25.7% and 22.9%, respectively). In the Task domain, attendance or availability-related responsibilities were the most common context (20.0%). In the Action domain, the most frequently reported behaviors were rule, ethics, and boundary violations (28.6%), followed by work avoidance or unavailability (17.1%) and inappropriate communication (17.1%). In the Result domain, the most common consequence was the impact on workflow or staff (20.0%), whereas direct patient impact was less frequent (5.7%) (Figure 1).

**Figure 1.**
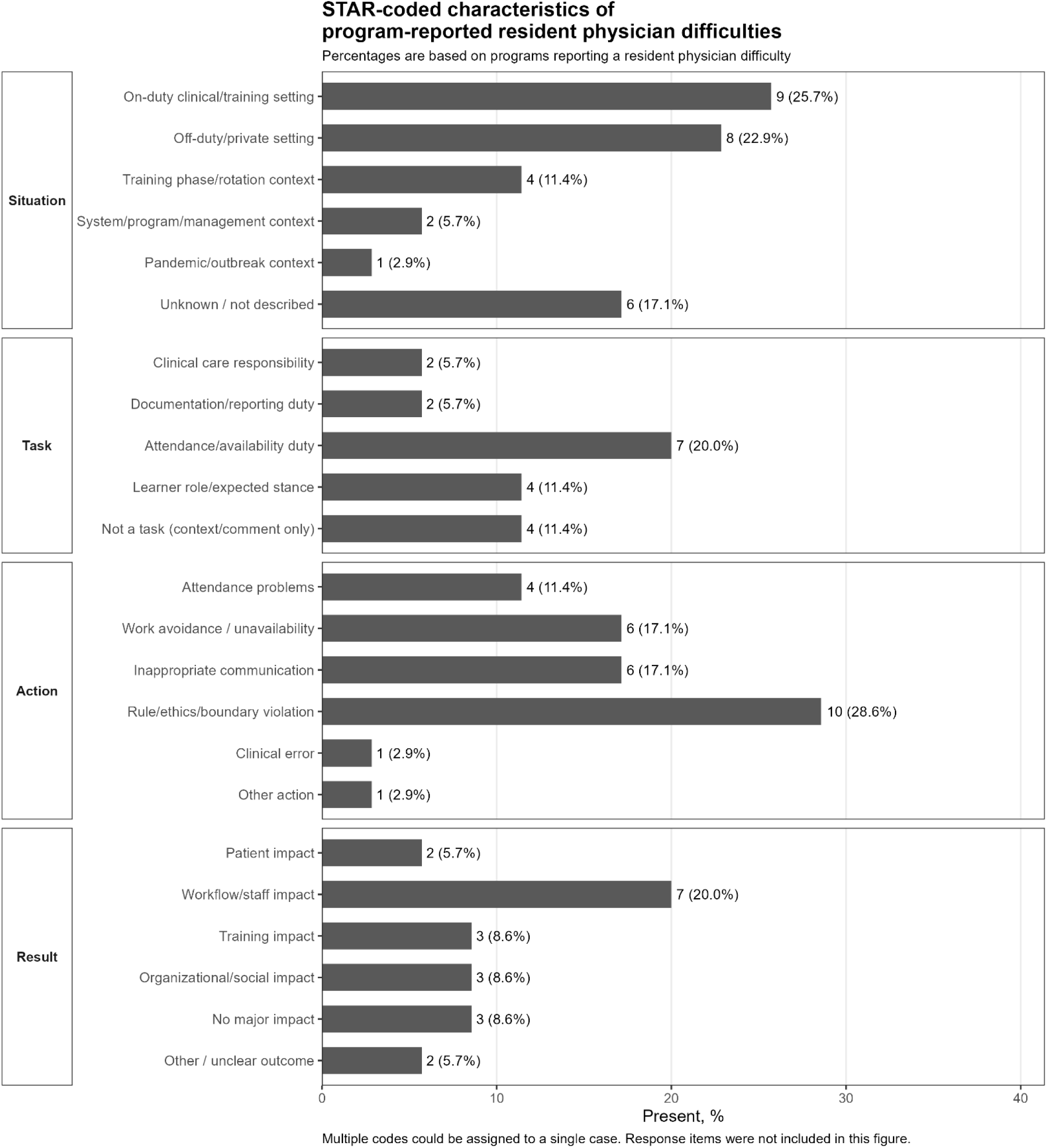
STAR-coded characteristics of program-reported resident physician difficulties. Note: Codes were derived from STAR-based free-text descriptions of program-reported resident physician difficulties. Multiple codes can be assigned to a single case; therefore, the percentages are not mutually exclusive. The program response codes are not included in the figure.

### Association between selection methods and resident physician difficulties

In analyses adjusted for hospital type and the number of GM-ITE examinees, the use of CBT scores was associated with the presence of resident physician difficulty (adjusted odds ratio [aOR], 4.60; 95% confidence interval [CI], 1.50–14.11; P = 0.008; FDR-adjusted P = 0.048). Essays, written academic tests, medical school grades, and aptitude test (personality assessment) scores were not significantly associated with the presence of a resident physician’s difficulty (Figure 2; Supplementary Table S1). The estimate for the aptitude test (ability assessment) is not plotted in Figure 2 because it was unstable owing to sparse data. Only three programs used this method, and no program-reported resident physician difficulty was observed among them (Supplementary Table S1). In the supplementary grouped analyses, programs that used the knowledge-based selection indicator did not have higher program-level GM-ITE total scores than those that did not use this indicator (Supplementary Table S2). In the supplementary item-level analyses, no selected non-interview selection method, including the use of the CBT score, was significantly associated with the program-level GM-ITE total score (Supplementary Table S4). In a supplementary grouped analysis, the knowledge-based selection indicator, defined as use of a written academic test conducted by the program and/or applicants’ pre-clinical-clerkship CBT scores as selection criteria, was not significantly associated with the presence of a program-reported resident physician difficulty after adjustment for hospital type and number of GM-ITE examinees (aOR, 1.74; 95% CI, 0.71–4.26; P = 0.225; Supplementary Table S3).

**Figure 2.**
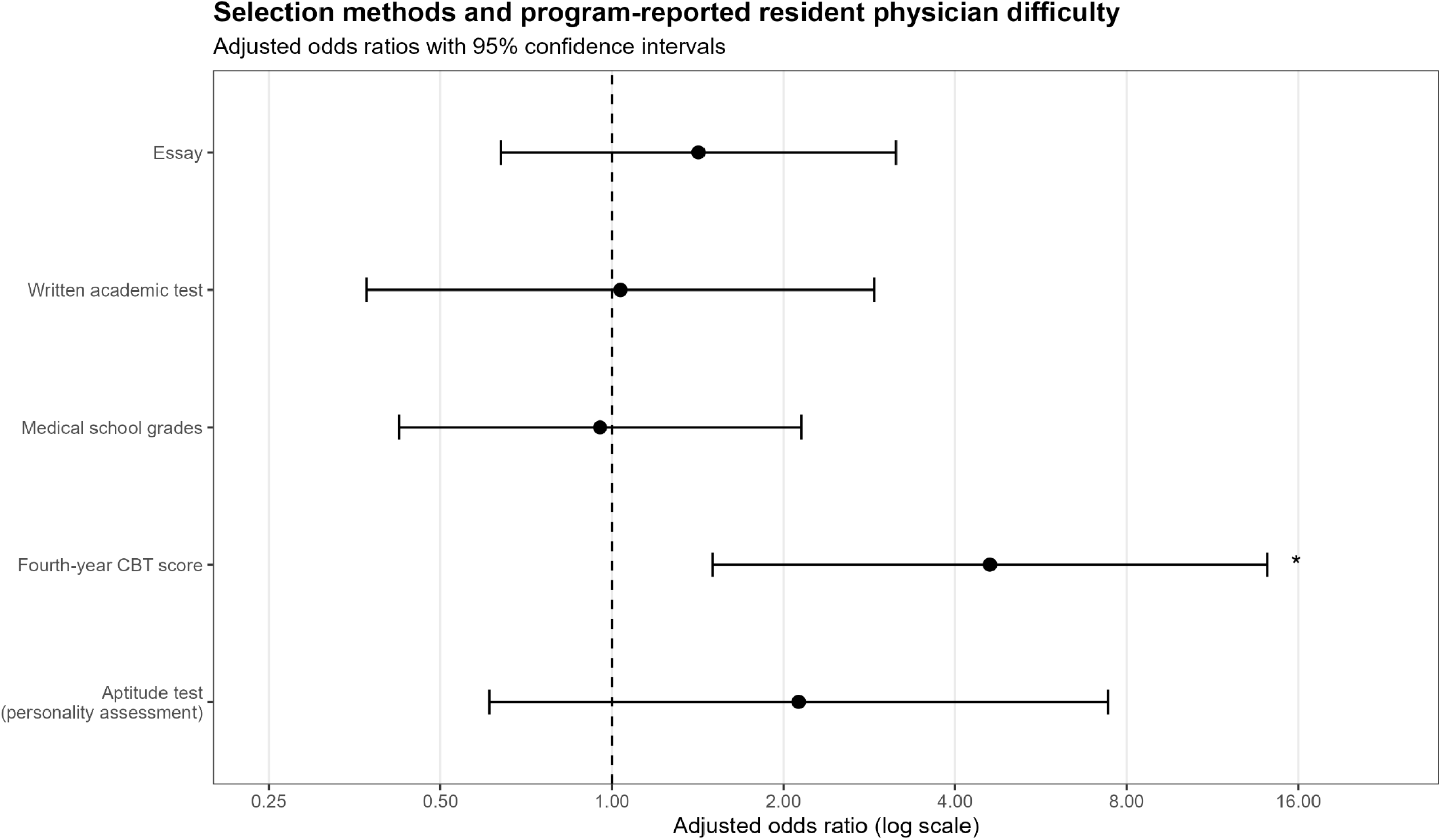
Adjusted associations between selected non-interview selection methods and the presence of a program-reported resident physician difficulty. Note: Odds ratios are estimated using separate multivariable logistic regression models for each selection method, adjusted for hospital type and number of GM-ITE examinees. Programs with missing responses for the resident physician difficulty items were excluded. The aptitude test (ability assessment) is not plotted because the estimate is unstable due to sparse data. Only three programs used this method, and no program-reported resident physician difficulties were observed among them. Asterisks indicate the Benjamini-Hochberg false-discovery rate-adjusted significance.

## Discussion

This cross-sectional study aimed to examine resident-physician selection practices in Japan, and whether the specific selection practices employed by each program were associated with program-reported resident physician difficulties and program-level GM-ITE performance. Our findings indicated that resident selection practices were centered on interviews, with variations in the use of other selection methods. Interviews were the predominant selection method and were also the most highly valued component of the selection process. Resident physician difficulties were reported by 35 of the 150 programs with valid responses. Among the selection methods examined, use of CBT score was associated with the presence of a resident physician difficulty, whereas other non-interview selection methods were not. Additionally, program-level GM-ITE performance did not differ significantly between programs with and without difficulty for resident physicians, and programs using knowledge-based testing did not show higher GM-ITE total scores.

To our knowledge, this is the first nationwide program-level survey to describe resident selection practices in Japan and to explore their associations with program-reported resident physician difficulties and program-level GM-ITE performance. In this study, program-reported resident physician difficulties were characterized primarily by issues related to professionalism and workplace conduct, including attendance problems, inappropriate communication, rules, ethics, and boundary violations. Moreover, these cases were more likely to disrupt workflow or affect staff than have direct effects on patients. These findings suggest that the reported difficulties of resident physicians were more often related to professionalism and workplace conduct than to direct clinical errors or patient harm.

Inappropriate communication was reported in some of the program-reported resident physician difficulties. Because communication is closely linked to professionalism in clinical practice, such difficulties may affect patient experience, treatment adherence, and broader clinical outcomes [16–22], while also contributing to professionalism-related concerns in team-based clinical settings. Work avoidance and unavailability were frequent patterns observed in our study. Such reliability-related problems may disrupt day-to-day workflow and team functioning, and unprofessional conduct can undermine team dynamics and psychological safety [23]. Although professionalism-related problems can ultimately affect patient care and safety, the immediate consequences reported in this study more commonly involve workflow or staff impacts than direct patient impacts.

In Japan, resident physicians rotate across multiple departments, often for relatively short periods [4]. One possible explanation is that, in this broad-rotation and supervised training structure, direct patient-level consequences may be less readily observed or attributed to individual residents through training programs, whereas disruptions in workflow, staff relationships, and supervision may be more visible.

Professionalism-related problems during residency have important downstream consequences, including remediation, disciplinary review, and disciplinary action [24,25]. Because such difficulties may become clearer through observed behavior during training, selection-stage indicators alone are unlikely to identify all residents who require support or remediation [26,27]. These findings reinforce the importance of identifying and addressing professionalism-related difficulties during training, rather than relying solely on selection stage indicators.

In our study, the use of CBT scores was associated with program-reported resident physician difficulties, whereas written academic tests and medical school grades were not. These indicators represent different types of academic information: medical school grades reflect institution-based evaluations during medical school, written academic tests are program-administered selection examinations, and CBT scores represent standardized pre-clinical clerkship knowledge assessments used as selection criteria. However, when written academic tests and CBT scores were combined into a broader knowledge-based testing category, no significant association was observed with program-reported difficulties of resident physicians. Furthermore, neither the CBT score nor the broader knowledge-based testing category was associated with higher program-level GM-ITE total scores. This finding suggests that knowledge-based indicators used during selection do not necessarily translate into higher program-level knowledge performance during residency. Therefore, knowledge-based indicators should be viewed as a component of readiness rather than a proxy for overall excellence as a resident. This pattern suggests that the association observed for the CBT score was specific to this item, rather than a general association between knowledge-based selection and resident physician difficulties. Concurrently, because the reported difficulties were mainly related to attendance, communication, and professionalism rather than knowledge deficits, adding knowledge-based indicators to an interview-centered selection process may not be sufficient to capture professionalism-related risk in its current form. This pattern fits within the broader international challenge of residency selection: programs must select applicants who are academically prepared for clinical training while also identifying attributes related to professionalism, communication, reliability, and fit with team-based practice. Internationally, residency selection commonly combines academic screening, examinations, letters, interviews, and ranking, with varying weights across countries and specialties [5–7]. However, professionalism-related outcomes are difficult to predict using selection data alone. Medical school professionalism concerns have been associated with later disciplinary action; however, their positive predictive value is limited [28–30]. Evidence from other settings also suggests that traditional academic metrics such as United States Medical Licensing Examination scores have limited utility in predicting professionalism-related outcomes, whereas behavioral or contextual information such as comments in dean’s letter, recommendation letters, interview flags, and structured professionalism-focused assessments may provide additional but imperfect signals [31–34]. Thus, the challenge is not simply whether to prioritize academic indicators or interviews but how multiple sources of information can be structured and interpreted to support fairer and more informative selection decisions.

These findings may also explain why interviews remain central to residency selection in Japan. In our study, interviews were not only the most commonly used selection method but also the most highly valued component of the selection process. The content of the important interview questions focused largely on why the applicant chose the hospital or program, career plans, and program fit, while measures to ensure interview quality relied mainly on multiple-interviewer systems and, to a lesser extent, on the inclusion of non-physicians. By contrast, explicitly structured interviews and scoring or quantification are uncommon. This pattern suggests that programs attempt to assess motivation, fit, and professionalism-related qualities through interviews, but these assessments are often conducted in relatively non-standardized ways.

Overall, these findings point to the limitations of relying on academic screening alone and raise the broader question of what qualities define an “excellent” applicant. Although knowledge-based indicators are important, they capture only one dimension of readiness for residency. Knowledge and test performance may change through clinical exposure, self-directed learning, supervision, and the training environment [35–37], whereas professionalism requires longitudinal development through role modeling, feedback, reflection, and remediation [38,39]. In contemporary clinical practice, where information is increasingly accessible and care is delivered through teams, excellence may depend on professionalism, communication, reliability, adaptability, and the ability to learn from feedback in addition to knowledge acquisition. Therefore, a more practical approach would be to combine careful selection with early, explicit, and longitudinal professionalism education. Professionalism concerns in medical students or early residents may be better understood as opportunities for feedback, support, and remediation rather than as fixed personal shortcomings [4,26–30].

This study has several limitations. First, although 151 residency programs participated, the findings may not be generalizable to all Japanese residency programs, because respondents may have differed from non-respondents in selection practices, reporting culture, or experience with program-reported resident physician difficulties. Second, self-reported data from program representatives may be subject to recall bias, underreporting, and differences in interpretation. Specifically, severe or sensitive cases may not have been completely reported. Third, the cross-sectional, program-level design precludes causal inference and does not allow the linkage of individual applicants’ selection data to subsequent behavior, remediation, or GM-ITE performance. Although we adjusted for the number of GM-ITE examinees as a proxy for the resident physician cohort size, this measure may not perfectly represent the exact number of PGY1 and PGY2 resident physicians or residency positions in each program, which may influence the probability of observing at least one reported resident physician difficulty.

Moreover, analyses involving the GM-ITE were conducted at the program level; therefore, program-level GM-ITE scores should not be interpreted as reflective of individual resident physicians’ performance after selection. These scores may reflect not only resident physicians’ baseline academic characteristics but also their clinical exposure, educational environment, supervision, and self-directed learning during residency. Thus, this study did not assess the individual-level predictive validity of CBT scores for subsequent GM-ITE performance. Fourth, the selection practice variables captured whether each method was used by the program, but not individual applicants’ or matched residents’ CBT scores, written examination scores, medical school grades, or the relative weights assigned to these components in the final selection decisions. Therefore, findings concerning knowledge-based testing represent program-level associations and do not directly evaluate individual-level test performance or the causal effects of specific selection tools. Finally, the definitions of program-reported resident physician difficulties and STAR-based descriptions depended on the program reports. The selection practices reported for 2023 may not be identical to those used when resident physicians involved in disciplinary actions or severe warnings are recruited. Moreover, the timeframe of the reported resident physician difficulties may vary across respondents. Therefore, the observed associations should not be interpreted as evidence that a specific selection method can cause or predict subsequent difficulties for resident physicians. Residual confounding and multiple-testing remain possible, and the findings are exploratory and hypothesis generating. This limitation also applies to the observed association between applicants’ pre-clinical clerkship CBT scores and program-reported resident physician difficulties, which may reflect program-level selection policies, applicant volume, reporting practices, or residual confounding factors.

## Conclusion

This nationwide program-level survey showed that resident selection in Japan remains strongly interview-centered, and program-reported resident physician difficulties were more often related to professionalism, communication, reliability, and workplace conduct than to knowledge deficits. The findings also suggest that academic selection indicators alone may be insufficient to identify applicants who may require additional support during residency. Future research should examine individual-level links between selection data and subsequent training outcomes and evaluate whether more structured interviews, explicit scoring criteria, and interviewer calibration improve the fairness and usefulness of resident selection.

## Data Availability

The data underlying this study are not publicly available because they contain potentially identifiable program-level information. De-identified data may be available from the corresponding author upon reasonable request, subject to approval by the relevant ethics committee and data custodian.

## Acknowledgments

We thank the program directors and supervising physicians who participated in this nationwide survey. We also thank the Japan Institute for Advancement of Medical Education Program for its support in administering the survey and managing the program-level data. We would like to thank Editage for English-language editing.

## Notes on contributors

Miwa Sekine, PhD, is affiliated with the Division of Medical Education, Juntendo University Faculty of Medicine, Tokyo, Japan.

Yuji Nishizaki, MD, MPH, PhD, is affiliated with the Division of Medical Education, Juntendo University Faculty of Medicine, and Clinical Translational Science, Juntendo University Graduate School of Medicine, Tokyo, Japan.

Takashi Watari, MD, is affiliated with the Integrated Clinical Education Center, Kyoto University Hospital, Kyoto, Japan.

Kiyoshi Shikino, MD, MHPE, PhD, AFAMEE, is affiliated with the Department of Community-Oriented Medical Education, Chiba University Graduate School of Medicine, Chiba, Japan.

Sho Fukui, MD, is affiliated with the Department of Emergency and General Medicine, Kyorin University, Tokyo, Japan.

Kazuya Nagasaki, MD, is affiliated with the Department of General Medicine, Fujita Health University, Aichi, Japan.

Masanori Nojima, MD, is affiliated with the Center for Translational Research, The Institute of Medical Science, The University of Tokyo, Tokyo, Japan.

Taro Shimizu, MD, is affiliated with the Department of Diagnostic and Generalist Medicine, Dokkyo Medical University Hospital, Tochigi, Japan.

Yu Yamamoto, MD, is affiliated with the Division of General Medicine, Center for Community Medicine, Jichi Medical University, Tochigi, Japan.

Hiroyuki Kobayashi, MD, is affiliated with the Department of Internal Medicine, Mito Kyodo General Hospital, University of Tsukuba, Ibaraki, Japan.

Yasuharu Tokuda, MD, is affiliated with the Muribushi Okinawa Center for Teaching Hospitals, Okinawa, Japan, and the Tokyo Foundation for Policy Research, Tokyo, Japan.

